# Interaural place-of-stimulation mismatch estimates using CT scans and binaural perception, but not pitch, are consistent in cochlear-implant users

**DOI:** 10.1101/2021.02.19.21251930

**Authors:** Joshua G. W. Bernstein, Kenneth K. Jensen, Olga A. Stakhovskaya, Jack H. Noble, Michael Hoa, H. Jeffery Kim, Robert Shih, Elizabeth Kolberg, Miranda Cleary, Matthew J. Goupell

## Abstract

ABSTRACT

Bilateral cochlear implants (BI-CIs) or a CI for single-sided deafness (SSD; one normally functioning acoustic ear) can partially restore spatial-hearing abilities including sound localization and speech understanding when there are competing sounds. However for these populations, frequency information is not explicitly aligned across the ears, resulting in interaural place-of-stimulation mismatch. This diminishes spatial-hearing abilities because binaural encoding occurs in interaurally frequency-matched neurons. This study examined whether plasticity – the reorganization of central neural pathways over time – can compensate for peripheral interaural place mismatch. We hypothesized differential plasticity across two systems: none for binaural processing but adaptation toward the frequencies delivered by the specific electrodes for sequential pitch perception. Interaural place mismatch was evaluated in 43 human subjects (20 BI-CI and 23 SSD-CI, both sexes) using interaural-time-difference (ITD) discrimination (simultaneous bilateral stimulation), place-pitch ranking (sequential bilateral stimulation), and physical electrode-location estimates from computed-tomography (CT) scans. On average, CT scans revealed relatively little BI-CI interaural place mismatch (26° insertion-angle mismatch), but relatively large SSD-CI mismatch, particularly at the apical end of the array (166° for an electrode tuned to 300 Hz, decreasing to 14° at 7000 Hz). ITD and CT measurements were in agreement, suggesting little binaural-system plasticity to mismatch. The pitch measurements did not agree with the binaural and CT measurements, suggesting plasticity for pitch encoding or procedural biases. The combined results show that binaural processing may be optimized by using CT-scan information, but not pitch measurements, to program the CI frequency allocation to reduce interaural place mismatch.

**SIGNIFICANCE STATEMENT:** Placement of electrode arrays in users of cochlear implants (CIs; bionic auditory prostheses that partially restore hearing) does not align the frequency information to acoustic neural encoding across the ears. This interaural place-of-stimulation mismatch diminishes spatial hearing abilities. This study shows that for experienced adult CI users with two CIs or with one CI and one normal-hearing ear, the best possible binaural sensitivity occurs when the same cochlear location is stimulated in both ears. This means that binaural brainstem pathways do not experience “plasticity” to compensate for interaural place mismatch – i.e., they do not reorganize to respond to input from different cochlear places. Therefore, explicit correction of interaural place mismatch by a clinician is necessary to derive maximum spatial-hearing benefits.

## Introduction

Neural computations in the superior olivary complex (SOC; Yin et al., 2019) underlie the auditory brainstem’s exquisite sensitivity to interaural differences in arrival time and intensity for a given sound source (Yost and Dye, 1988; Thavam and Dietz, 2019), providing tremendous functional advantages for sound-source localization and speech understanding in noise (Reeder et al., 2014). In an attempt to provide some of these advantages to severely hearing-impaired individuals, there are a growing number of cochlear-implant (CI) users with two functional ears through bilateral CIs (BI-CIs; for people with bilateral severe-to-profound loss; Peters et al., 2010) or one CI for people with single-sided deafness (SSD-CI; normal/near-normal acoustic hearing in the contralateral ear; Buss et al., 2018).

While BI-CI and SSD-CI users experience some functional advantages of two ears (Litovsky et al., 2012; Bernstein et al., 2016, 2017), binaural benefit is diminished compared to normal-hearing individuals. Various factors inherent to CI processing might limit binaural benefits, such as lack of temporal fine-structure encoding (Churchill et al., 2014) or a time delay between the CI and acoustic ears (Zirn et al., 2015). Yet many BI- CI and SSD-CI users show binaural sensitivity under laboratory conditions (Kan and Litovsky, 2015; Bernstein et al., 2018), corroborating the evidence for binaurally sensitive brainstem pathways with electric stimulation (Chung et al., 2016).

Binaural sensitivity in the SOC depends on the symmetric tonotopicity of the auditory periphery (Blanks et al., 2007) and interaural place-of-stimulation mismatch reduces binaural sensitivity (Poon et al., 2009; Kan et al., 2015). While rarely a problem in acoustic hearing, the cochlear place of electrical stimulation is determined by physical electrode location. Thus, many BI-CI and most SSD-CI users have frequency- mismatched ears (Kawano et al., 1998; Aschendorff et al., 2005; Landsberger et al., 2015), which can reduce binaural benefit for speech understanding (Wess et al., 2017; Sagi et al., 2020) and localization (Suneel et al., 2017). In normal-hearing systems, SOC neurons are tuned to respond to binaural inputs from matched cochlear places. A critical question is if binaural neural pathways exhibit “plasticity” to overcome interaural place mismatch: Can binaural neurons adapt to respond to input from interaurally different cochlear locations? CI users show evidence of plasticity to frequency-to-place mismatch to better understand speech in one ear (Svirsky et al., 2004) and to reduce interaural differences in place-pitch perception (Reiss et al. 2007, 2014). Functional plasticity has been observed for changes in the mapping of binaural cues to a physical sound-source location (Van Wanrooij and Van Opstal, 2005). However, in each of these cases, the observed changes might only reflect a relearning of cue meaning, without requiring brainstem rewiring.

Hu and Dietz (2015) addressed the question of whether the binaural system exhibits plasticity to overcome interaural place mismatch by comparing binaural sensitivity (using electrophysiological and perceptual measures) to sequential pitch comparisons for BI-CI users. While an electrode in one ear was usually pitch-matched to the same-numbered electrode in the other, binaural sensitivity was often optimal for different- numbered electrodes. This was interpreted as evidence for plasticity for place-pitch perception but not binaural processing. The evidence, however, was indirect because physical electrode locations were unknown.

To address the question of plasticity to interaural place mismatch more directly, the current study explicitly compared interaural place-pitch perception and tuning curves for interaural time-difference (ITD) sensitivity to estimates of electrode position derived from computed-tomography (CT) scans. Previous studies have compared pitch matching and ITD discrimination for BI-CI users (Hu and Dietz, 2015), or pitch matching and CT scans for SSD-CI users (Schatzer et al., 2014; Adel et al., 2019), but the three measurements have never been compared. We hypothesized that evidence for plasticity would differ between the two perceptual measures: binaural sensitivity would not show evidence of plasticity, with ITD-based estimates aligning closely with CT-based estimates of electrode location, while interaural place pitch would show evidence of plasticity, with pitch-based estimates aligning more closely with clinical center frequencies (CFs) in the CI map.

## Materials and Methods

### Subjects

A total of 43 subjects (20 BI-CI and 23 SSD-CI) participated in the study (Tables I and II). These subjects had a wide range of ages, duration of deafness, and etiology, but all subjects had used their CIs for at least 6 months at the time of the study. The BI-CI subjects were all implanted in both ears with Cochlear Ltd. (Sydney, Australia) devices. SSD-CI subjects were implanted with a Cochlear Ltd. or MED-EL (Innsbruck, Austria) device in one ear, while the other ear had either normal hearing thresholds (N=16; ≤25 dB HL for octave frequencies between 250-4000 Hz) or mild (N=5; ≤40 dB HL) or moderate hearing loss (N=2; ≤60 dB HL). Four subjects with mild or moderate hearing loss usually wore a hearing aid in everyday listening situations, but did not use the hearing aid in the experiments described here. Testing was performed at the University of Maryland-College Park and Walter Reed National Military Medical Center in a quiet sound booth. The Institutional Review Boards at each institution approved this research protocol. Informed consent was obtained from subjects before testing.

**Table I.** Demographic and device information for the BI-CI subjects.

| Subject | Age | Sex | Left ear |  |  | Right ear |  |  |
| --- | --- | --- | --- | --- | --- | --- | --- | --- |
|  |  |  | Duration of<br>deafness<br>(range, yrs) | CI<br>experience<br>(range, yrs) | Internal<br>Device | Duration of<br>Deafness<br>(range, yrs) | CI<br>experience<br>(range, yrs) | Internal<br>Device |
| BCI1 | 70s | F | 31-40 | 21-30 | CI24M | 41-50 | 10-15 | CI24R(CS) |
| BCI2 | 70s | M | 3-5 | 15-20 | CI24R(CS) | 6-10 | 6-10 | CI24RE |
| BCI3 | 60s | F | 1-2 | 11-15 | CI24RE | 1-2 | 11-15 | CI24RE |
| BCI4 | 70s | F | <1 | 15-20 | CI24R(CS) | 3-5 | 11-15 | CI24RE |
| BCI5 | 60s | F | 3-5 | 11-15 | CI24RE | 6-10 | 11-15 | CI24R(CA) |
| BCI6 | 70s | M | 21-30 | 3-5 | CI422 | <1 | 11-15 | CI24R(CS) |
| BCI7 | 50s | F | 11-15 | 6-10 | CI24RE | 11-15 | 6-10 | CI24RE |
| BCI8 | 60s | F | 11-15 | 6-10 | CI512 | 11-15 | 6-10 | CI24RE |
| BCI9 | 70s | M | 61-70 | 3-5 | CI24RE | 1-2 | 6-10 | CI512 |
| BCI10 | 50s | F | <1 | 6-10 | CI512 | 1-2 | 3-5 | CI512 |
| BCI11 | 70s | F | 11-15 | 6-10 | CI24RE | <1 | 11-15 | CI24RE |
| BCI12 | 70s | F | 1-2 | 6-10 | CI24RE | 1-2 | 6-10 | CI512 |
| BCI13 | 60s | M | 11-15 | 6-10 | CI24RE | 21-30 | 3-5 | CI24RE |
| BCI14 | 60s | F | 6-10 | 6-10 | CI512 | 6-10 | 6-10 | CI512 |
| BCI15 | 70s | F | 21-30 | 6-10 | CI24RE | 21-30 | 6-10 | CI512 |
| BCI16 | 60s | F | 1-2 | 11-15 | CI24R(CS) | 6-10 | 3-5 | CI24RE |
| BCI17 | 60s | F | <1 | 6-10 | CI24RE | <1 | 6-10 | CI512 |
| BCI18 | 50s | F | 21-30 | 3-5 | CI422 | 15-20 | 6-10 | CI24RE |
| BCI19 | 40s | M | <1 | 1-2 | CI532 | 6-10 | 3-5 | CI24RE |
| BCI20 | 20s | M | <1 | 6-10 | CI24RE | <1 | 6-10 | CI24RE |

**Table II.**
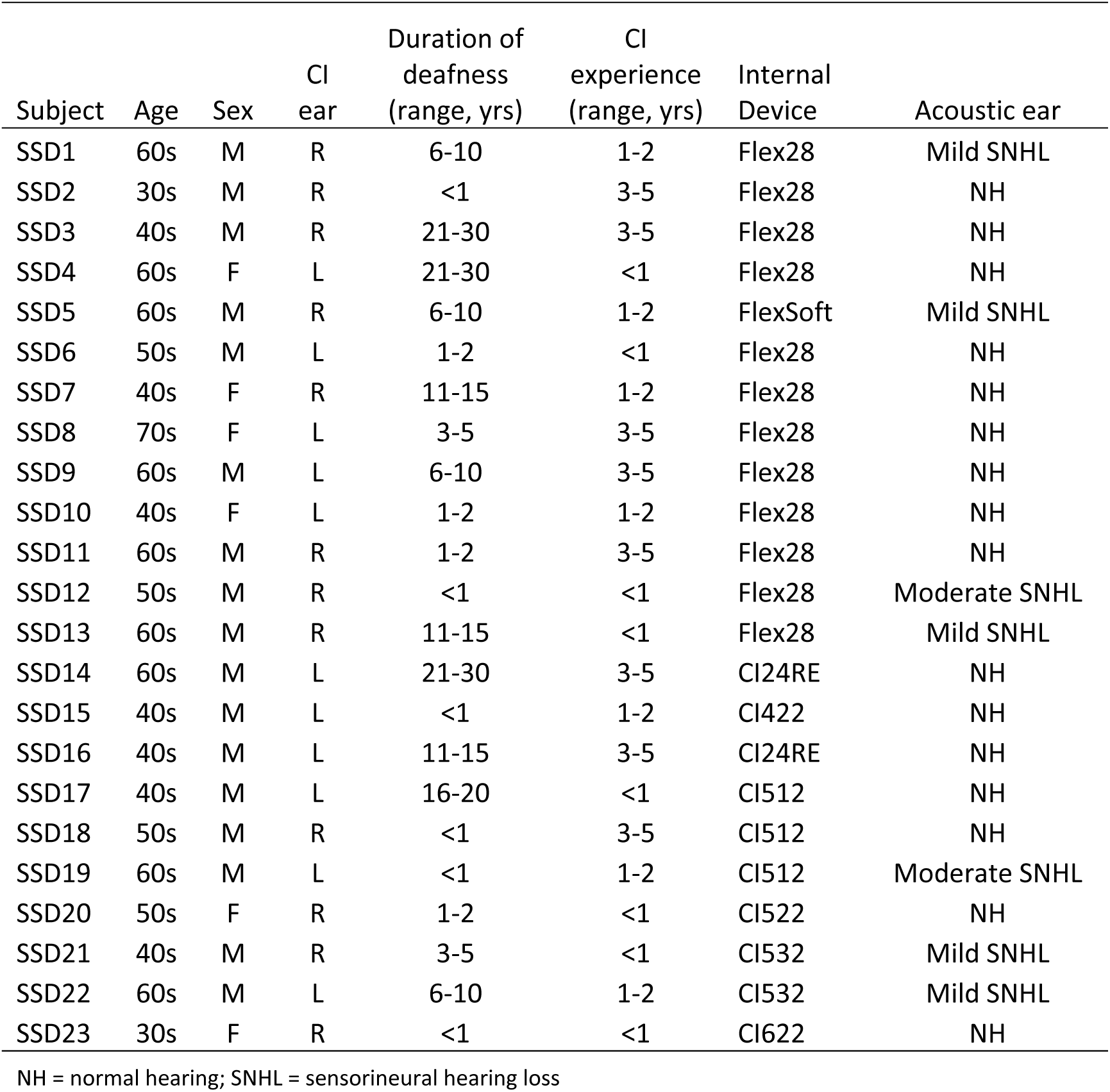
Demographic and device information for the SSD-CI subjects.

| Subject | Age | Sex | CI ear | Duration of deafness (range, yrs) | CI experience (range, yrs) | Internal Device | Acoustic ear |
| --- | --- | --- | --- | --- | --- | --- | --- |
| SSD1 | 60s | M | R | 6-10 | 1-2 | Flex28 | Mild SNHL |
| SSD2 | 30s | M | R | <1 | 3-5 | Flex28 | NH |
| SSD3 | 40s | M | R | 21-30 | 3-5 | Flex28 | NH |
| SSD4 | 60s | F | L | 21-30 | <1 | Flex28 | NH |
| SSD5 | 60s | M | R | 6-10 | 1-2 | FlexSoft | Mild SNHL |
| SSD6 | 50s | M | L | 1-2 | <1 | Flex28 | NH |
| SSD7 | 40s | F | R | 11-15 | 1-2 | Flex28 | NH |
| SSD8 | 70s | F | L | 3-5 | 3-5 | Flex28 | NH |
| SSD9 | 60s | M | L | 6-10 | 3-5 | Flex28 | NH |
| SSD10 | 40s | F | L | 1-2 | 1-2 | Flex28 | NH |
| SSD11 | 60s | M | R | 1-2 | 3-5 | Flex28 | NH |
| SSD12 | 50s | M | R | <1 | <1 | Flex28 | Moderate SNHL |
| SSD13 | 60s | M | R | 11-15 | <1 | Flex28 | Mild SNHL |
| SSD14 | 60s | M | L | 21-30 | 3-5 | CI24RE | NH |
| SSD15 | 40s | M | L | <1 | 1-2 | CI422 | NH |
| SSD16 | 40s | M | L | 11-15 | 3-5 | CI24RE | NH |
| SSD17 | 40s | M | L | 16-20 | <1 | CI512 | NH |
| SSD18 | 50s | M | R | <1 | 3-5 | CI512 | NH |
| SSD19 | 60s | M | L | <1 | 1-2 | CI512 | Moderate SNHL |
| SSD20 | 50s | F | R | 1-2 | <1 | CI522 | NH |
| SSD21 | 40s | M | R | 3-5 | <1 | CI532 | Mild SNHL |
| SSD22 | 60s | M | L | 6-10 | 1-2 | CI532 | Mild SNHL |
| SSD23 | 30s | F | R | <1 | <1 | CI622 | NH |

NH = normal hearing; SNHL = sensorineural hearing loss

### Interaural time-difference discrimination

The ITD discrimination test used electric pulse trains delivered to a single CI electrode, or bandlimited acoustic pulse trains delivered to an acoustic-hearing ear, to measure an ITD sensitivity tuning curve. The general methodology was to present a pulse train to one fixed electrode in the *reference* ear, and to measure ITD sensitivity as a function of electrode number (BI-CI subjects) or acoustic carrier frequency (SSD-CI subjects) in the other *comparison* ear. Interaural place mismatch was then estimated by determining the comparison electrode or carrier frequency that yielded the best ITD sensitivity. Within this general framework, it was necessary to use different methodologies for the ITD sensitivity task across the two subject groups.

#### BI-CI subjects

The BI-CI subjects performed a two-interval, two-alternative forced-choice task that generally followed Kan et al. (2015). Sequential pairs of binaurally presented equal-amplitude pulse trains were presented. The two stimuli had the same ITD magnitude but in opposite directions, and subjects were asked to determine whether the stimulus was perceived as moving from left to right or from right to left.

For most subjects and reference electrodes, an adaptive procedure was used to estimate the smallest detectable change (just-noticeable difference, JND) in ITD. JNDs were defined based on the relative ITD between the two intervals in the task (i.e., a 5000-µs ITD represents a condition where the ITD was +2500 µs in one interval and −2500 µs in the other). For a given reference/comparison electrode pair, the starting ITD was 5000 µs. The ITD magnitude then changed from trial-to-trial, following a three-down, one-up tracking rule (Zwislocki and Relkin, 2001) to estimate the 75% correct point on the psychometric function. The ITD changed by a factor 2 until the second reversal, a factor of 1.414 until the fourth reversal, and a factor of 1.2 thereafter, with the ITD values rounded to the nearest 20 µs. For ITDs below 200 µs, the adjustment step size was fixed at 20 us. The track continued for a total of ten reversals, and the JND was estimated as the geometric mean of the ITD for the last six reversal points. At least three adaptive tracks were completed for each combination of reference and comparison electrode, except when time constraints allowed only one (5%) or two runs (5% of cases). In cases where the standard deviation of the ITD JND was >25%, we added 2 more tracks, time permitting.

For a small number of initial cases (three subjects and two reference electrodes each, constituting approximately 5% of the data collected) a method of constant stimuli was used to estimate ITD JNDs, although we moved away from this initial this approach because it was too time consuming. For these measurements, ideally 50 trials were presented for each of a range of ITDs that produced a well-defined psychometric function, typically four points covering the range from near-chance to near-perfect performance. ITD values typically started as 200, 400, 800, 1600 µs, but then adjusted between 20 µs and 4000 µs depending on the sensitivity of the subject. Fewer trials (but always at least 36 per condition) were sometimes necessary given time constraints. Psychometric functions were fit to the data describing the percentage correct as a function of ITD, and the JND was estimated based on the 75% correct point on this function.

For each subject, one ear was designated as the reference ear. In most cases, the reference ear was selected to be the ear with the poorer speech-reception performance in quiet. In cases were monaural speech reception scores were similar between the ears, the subject’s non-preferred ear was selected as the reference ear. This was done with the idea that the mismatch data collected might ultimately be used to modify the clinical CFs in the reference ear to reduce interaural place mismatch. Modifying the poorer ear would avoid disrupting speech cues in the ear that a listener generally relies on more heavily.

Each subject performed a series of ITD discrimination estimates for up to five electrodes in the reference ear (time-permitting). For most subjects, the five electrodes chosen (out of the 22 electrodes in the array) were E4 (near the high-frequency, basal end of the array), E8, E12, E16, and E20 (near the low-frequency, apical end of the array. In some cases, where a subject had one of these electrodes deactivated (e.g., extra cochlear, facial nerve stimulation, or unpleasant auditory sensation), a different nearby reference electrode was selected. For a given reference electrode, the range of comparison electrodes was initially selected to include every even-numbered electrode within ±8 electrodes of the number-matched equivalent (e.g., for reference electrode E10, the range of comparison electrodes was E2–E18; for E4, the range was E2-E12). Additional even-numbered comparison electrodes and measurements were added until the subject could not reliably perform the task (>4 incorrect responses in a single block for an ITD of 5000 µs).

Electric stimulation was presented using direct-stimulation research equipment and software, consisting of a pair of Nucleus Freedom programming pods, L34 sound processors, and NIC2 software (Cochlear Ltd., Sydney, Australia) controlled by custom MATLAB scripts (Mathworks, Natick, MA). Most subjects were presented constant-amplitude, 100- or 200-pulses-per-second (pps) monopolar pulse trains to one electrode in each ear, using biphasic pulses that were 58 µs in duration (25 µs anodic and cathodic phases separated by an 8 µs gap). Synchronization was achieved via a trigger signal delivered from one pod to the other.

Prior to the experiment, loudness balancing was carried out to assure that stimulation was presented at a comparable loudness level for each electrode. The loudness-balancing procedure was carried out separately for the left and right ears. First, subjects were asked to identify a comfortable loudness level for each individual electrode stimulated in isolation, while ignoring the pitch of the stimulation. Second, subjects were presented with stimulation on five sequential electrodes and asked to judge whether there were any notable loudness differences between the five stimuli. Adjustments were made to the levels of the individual electrodes, and the five-electrode sweep repeated, until the subject reported that the five electrodes were comparable in loudness. Then this process was repeated with different selections of five electrodes until all available electrodes were exhausted. Finally, small adjustments were made to the levels of the reference electrodes to intracranially center the sound image for each number-matched pair, while keeping the non-reference-ear electrodes loudness balanced within that ear.

#### SSD-CI subjects

For the SSD-CI subjects, the procedure was as described Bernstein et al. (2018). For these subjects, the fact that one ear was electrically stimulated while the other was acoustically stimulated meant that there was an unknown relative delay in the time between stimulus presentation and auditory-nerve response in each ear (Zirn et al., 2015). This made it difficult to create stimuli with equal but opposite effective ITDs as was done for the BI-CI subjects, thus preventing this approach. While procedures have been developed using auditory brainstem responses (Zirn et al., 2015) or psychophysical image centering (Francart et al., 2018) to determine this relative delay, the amount of time needed for these procedures was not feasible for the purposes of the current study because of the need to find this point for numerous reference-comparison locations. Thus, instead of requiring subjects to discriminate the perceived direction of an interaurally delayed stimulus, the current study asked subjects to detect a change in ITD in a three-interval, two-alternative forced-choice task.

On each trial, subjects were presented with three intervals, with each interval containing four binaurally presented pulse-train bursts. In each of the two reference intervals, the four bursts all had the same ITD. In the target interval, the ITD varied between the four bursts. Subjects were instructed to identify which of the three intervals contained the stimulus that was “moving” or “changing”. The first interval was always the reference, and the subject was required to identify whether the target stimulus occurred in the second or third interval. The ITDs for the four bursts in the target interval were evenly spaced and symmetrically placed around the fixed ITD in the reference interval. For example, if the reference interval ITD was fixed at 500 µs for all four bursts, and the ITD spacing between bursts in the target interval was 2500 µs, then the four target-interval ITDs were −3250, −750, 1750, and 4250 µs. The pulse rate was in most cases 100 pps. For some reference electrodes for some subjects the pulse rate was instead set to 50 pps, either because the subject was unable to detect ITD changes at 100 pps, or because acoustic carrier frequencies below 1000 Hz were being tested (see below).

Electrical stimulation was delivered to the desired CI electrode using one of two methods depending on the CI manufacturer. For the SSD-CI subjects who used Cochlear Ltd. devices, the same direct-stimulation interface described above for the BI-CI subjects was employed. The only difference was that synchronization of the electric and acoustic channels was achieved by initiating electrical stimulation via a trigger delivered from the second channel of the RME Hammerfall Multiface II sound card (RME, Haimhausen, Germany) that was used to generate the acoustic stimulus in the other ear. The trigger signal was amplified (Tucker-Davis Technologies, Alachua, FL) before being delivered to the programming pod. Interaural timing was calibrated by time-aligning the center of the acoustic pulse (at the headphone transducer) to the center of the electric pulse at the electrode, as measured using a Cochlear Freedom Implant Emulator (Cochlear Ltd., Sydney, Australia) containing a CI internal device and resistive load that approximated the impedance inside the human cochlea.

For the SSD-CI subjects who used MED-EL devices, electrical stimulation was delivered via auxiliary input to an Opus 2 processor that was programmed with up to four different single-channel maps. Each single-channel map was created by reducing the C levels for all channels except for the stimulated electrode to zero, then setting the frequency cutoffs to the widest possible range allowed by the clinical software (5049– 8010 Hz). The stimulation electrode was then selected by choosing the appropriate program using the remote control. An acoustic pulse train (see below for details) with a center frequency of 6529 Hz and a bandwidth equivalent to 1.5 mm on the Greenwood (1990) scale, at the same pulse rate as the electrical stimulus in the other ear, was then delivered to the auxiliary input of the processor.

Acoustic stimulation was delivered via HD280 pro circumaural headphones (Sennheiser, Wedemark, Germany) to the desired cochlear place in the non-implanted ear also as described by Bernstein et al. (2018). The acoustic stimuli consisted of trains of Gaussian envelope pulses (Goupell et al., 2010, 2013), also known as Gabor (1947) pulses, constructed by modulating a sinusoidal carrier tone by a Gaussian shaped envelope. The acoustic pulses were presented at the same rate as the electrical pulses in the other ear and had a 3-dB bandwidth equivalent to 1.5 mm on the Greenwood scale. The acoustic carriers were selected from a fixed set of frequencies, defined in 1.5-mm steps on the Greenwood scale (473, 616, 791, 1007, 1272, 1598, 2000, 2494, 3102, 3850, 4770, 5901, 7294, 9007, and 11114 Hz).

The combination of pulse rates and carrier frequencies that could be reliably presented to subjects was limited by the physics of the stimulus generation and by the resolving capabilities of the cochlea. Because the bandwidth (in Hz) decreased with decreasing carrier frequency, this meant that the temporal width of the pulses become larger with decreasing frequency. In most cases, subjects were presented with 100-pps pulse trains. However, at this rate, successive pulses would have overlapped by more than 1% in linear amplitude for carrier frequencies below 1140 Hz (Goupell et al., 2010). Furthermore, for low carrier frequencies, frequency components below about the 10^th^ harmonic are resolved by the auditory system (Bernstein and Oxenham, 2003) and therefore do not interact in the cochlea to generate an envelope at the desired pulse rate. This means that for a pulse rate of 100 pps, the intended pulse-envelope would not be represented for frequencies below about 1000 Hz. Therefore, for relatively apical (i.e., low-frequency) reference electrodes where acoustic carrier frequencies ≤1000 Hz were included in the set of comparison stimuli, a pulse rate of 50 pps was used instead.

Subjects were generally tested on three different reference electrodes in the CI ear, although some subjects were tested on four or five electrodes when time permitted. For each reference electrode, ITD discrimination was measured for a range of at least five, and in most cases 6-10, carrier frequencies for the acoustic pulse trains delivered to the other ear, with at least 30 trials per condition. For each electrode, several parameters of the electric and acoustic pulse trains, and the range of acoustic carrier frequencies were adjusted in pilot tests, with the goal of identifying a stimulus that would yield a maximum level of performance in the range of 80-90% correct, which would be high enough to estimate a tuning function while avoiding ceiling effects. The parameters that were adjusted included: the reference interval ITD, the spacing between the ITDs in the target interval, the pulse rate, and the burst duration. Following Bernstein et al. (2018), the default values for these parameters depended on the CI manufacturer.

Prior to the experiment, loudness balancing was carried out for each of the reference electrodes and acoustic carrier frequencies tested in the study. Electric and acoustic stimuli were presented sequentially and the subject was asked to indicate which stimulus was perceived as louder. The experimenter then adjusted the level of the stimulus presented to one ear in steps of 1-3 dB (or in steps of 1-5 clinical current units for direct electrical stimulation) until the subject reported that the two stimuli were at equal loudness. For a given reference electrode, subjects were first presented with an acoustic pulse train with a carrier frequency of 2000 Hz and a level of 50-60 dB SPL, and the level of the electrical stimulus was adjusted to create a loudness match. Then, the electrical stimulus was held fixed at that level, and the levels for each individual acoustic carrier frequencies were adjusted to create a loudness match to the electrical stimulus. In cases where a loudness match could not be reached within the limits of the system, which sometimes occurred for the highest frequencies tested, the level of the electrical stimulus was reduced and the loudness balancing procedure repeated for that reference electrode.

#### Analysis

For both subject groups, the ITD tuning curves were fit with a skewed-Gaussian function with four free parameters describing: (1) the peak performance (best JND or percentage correct), (2) the width and (3) the skewness of the function (Bernstein et al., 2018), and (4) the comparison electrode or frequency where this peak occurred. This fourth quantity served as the estimate of the interaural match – i.e. the comparison ear electrode or acoustic frequency that yielded maximum performance. For the BI-CI subjects, chance performance for the purposes of curve fitting was taken to be the maximum allowed ITD in the adaptive track, which was either 2500 or 5000 us. For the SSD-CI subjects, chance performance for the purposes of curve fitting was taken to be 50%.

### Pitch Ranking

To estimate the perceived relative place pitch between the two ears, this experiment used the “mid-point comparison” pitch-ranking methodology (Long et al., 2005; Cosentino et al., 2016). This technique was developed as a fast way to pitch-rank a set of electrodes in the two ears, and was found by Jensen et al. (2021) to yield a set of place pitch estimates for BI-CI subjects that are relatively immune to procedural bias effects, including the influence of comparison-electrode test range (Carlyon et al., 2010; Goupell et al., 2019) and starting point on the pitch-match estimate. While there were some differences in how this methodology was carried out for the BI-CI versus SSD-CI subjects (see below), the algorithm was generally similar for the two groups. Subjects were asked to pitch rank a set of stimuli consisting of a subset of electrodes or acoustic pure tones from each ear. This was accomplished through a series of pairwise comparisons, where each “new” electrode or pure tone added to the ranking was compared to the existing ranking in an adaptive manner. First, the “new” electrode or pure tone (the reference stimulus) was compared in pitch to one of the already ranked electrodes or pure tones (the comparison stimuli). Based on the subject’s response regarding which of the reference or comparison stimulus was perceived to be higher in pitch, the possible choices for ranking position of the reference stimulus was reduced to only those electrodes/tones that were higher or lower in the pitch ranking than the previous comparison stimulus. The next comparison stimulus was then chosen to be the midpoint of this new reduced range. This process was repeated iteratively until the stimulus choices were exhausted such that the reference stimulus was assigned to a position in the ranking. Then the process was repeated with each new reference stimuli, until the entire stimuli were ranked. This ranking process was repeated 10 times for each subject.

#### BI-CI subjects

The BI-CI subjects pitch ranked all even-numbered electrodes from the two ears (up to a total of 22 electrodes, 11 for each ear). Single 300-ms bursts of a 1000-pps pulse train were presented to two successive electrodes with an inter-stimulus interval of 300 ms. In each block, the subject was initially presented with two sequential bursts on two different electrodes chosen at random, and their response regarding with electrode was higher established the rank order for these two electrodes. Then, a third electrode was compared to one of the two electrodes in the existing ranking, chosen at random, depending on the response, it might also have been compared to the other electrode in the existing ranking, to establish a ranking of the three electrodes. Subsequent electrodes were added into the ranking one at a time, as described above, until all 22 electrodes were rank ordered. This process repeated 10 times for each subject.

#### SSD-CI subjects

For the SSD-CI subjects, there were two considerations that required changes to the pitch-ranking protocol relative to the BI-CI subjects. The first consideration was that for the acoustic ear, the step sizes (1.5-mm spacing between adjacent stimulus frequencies) were so large relative to frequency-discrimination thresholds that subjects could rank order these pure-tones with nearly perfect accuracy. Therefore, the acoustic tone frequencies were assumed to be perfectly rank ordered from the beginning, and individual CI electrodes were added into this rank ordering. The second consideration was that for the MED-EL subjects, the electrical stimulation on a given electrode was generated by presenting a pure-tone stimulus to the auxiliary input of the sound processor loaded with single-channel maps. Because the intended electrode was selected manually by changing the program with the remote control, and only four single-channel maps could be stored in a given processor, it was not possible to complete a full ranking of the electrodes and acoustic frequencies.

To keep the procedure as similar as possible between the Cochlear and MED-EL users, all the SSD-CI subjects were tested in a modified pitch-ranking methodology where only one electrode at a time was ranked relative to the fixed set of acoustic frequencies. For the MED-EL subjects, up to seven different electrodes were tested using this methodology by loading four single-electrode maps to one sound processor and another set of three single-electrode maps to a second sound processor. For the Cochlear subjects, up to 11 different electrodes were tested using direct stimulation. Five pitch-ranking blocks were completed for each single electrode before changing the maps to test a different electrode. This process was completed twice, for a total of 10 pitch-ranking blocks per electrode.

#### Analysis

The idea was that in any future remapping that would be done to reduce interaural mismatch, only the clinical map CFs for the poorer ear would be adjusted, to avoid making changes to the ear upon which subjects rely on more heavily. Therefore, to identify the frequency range to which a given poorer-ear (reference) electrode should be mapped, the relevant information was to identify the better-ear (comparison) electrode number or acoustic frequency that was best pitch-matched to the poorer-ear electrode.

For the BI-CI subjects, the data were analyzed by plotting the average rank (among the 22 electrodes tested) for each of the 11 electrodes tested in each ear, then fitting the data for each ear with a sigmoidal function. For a given poorer-ear electrode, the matching better-ear electrode was identified by extracting from these fitted functions the better-ear electrode that yielded the same average rank.

For the SSD-CI subjects, the pitch-ranking data were analyzed to determine which pure-tone frequency was most closely matched to a given CI electrode. Because the acoustic pure tones were assumed to be correctly rank ordered, fitting was not required to extract this information from the ranking data. For each pitch-ranking block, the best-matching frequency was defined to be the midpoint (on the Greenwood scale) between the two frequencies above and below the CI electrode in the ranking. The mean matched frequency was then determined by averaging (on the Greenwood scale) the 10 blocks for each electrode.

#### Bias checks

Carlyon et al. (2010) argued that interaural pitch-match estimates can be influenced by the parameters of the available comparison-ear stimuli rather than the perceived pitch of the electrode of interest in the reference ear. They proposed checks that could be carried out to verify a minimal effect of such influences. For an adaptive pitch-matching procedure, they proposed that a pitch match can be considered valid if it is relatively immune to changes in the starting point of the adaptive track. Specifically, they proposed a criterion ratio of 0.5: for a given change in the starting point of the adaptive track, a pitch match is considered valid if it changes by less than half that amount. For each electrode tested, the slope of the relationship between the pitch rank and the adaptive-track starting point was calculated. For the BI-CI subjects, the mean slope was −0.03 (± 0.33 SD) and 4.7% of electrodes tested had slope greater than 0.5. For the SSD-CI subjects, the mean slope was 0.05±0.17 and 1.7% of electrodes tested had slope greater than 0.5. Thus, nearly all of the pitch matches passed the starting-point bias check.

### Computed Tomography Scans

Cochlear implant CT scans were acquired on a multidetector row CT scanner with a special temporal bone protocol that included extended Hounsfield unit (HU) scale implementation. Other scan parameters were 0.6-mm collimation, 140-kVp tube voltage, 300-mAs tube current (without modulation), 0.3-mm spacing between slides, and bone kernel for image reconstruction. In addition to standard temporal bone images in the axial and coronal planes, a 10 cm field-of-view oblique Stenver reformat parallel to the basal turn of the cochlea was created for better depiction of the implant electrode array position with ultra-high in-plane resolution of 0.2 × 0.2 mm.

#### Analysis

An automated image analysis sequence was used to determine the intra- cochlear location of the electrodes. A so-called “statistical shape model” (Cootes et al., 1995) of intracochlear anatomy based on the manual delineation of structures in micro-CT scans from 16 cadaveric specimens was used to determine patient-specific cochlea shape from CT scans (Noble et al., 2011). The model encodes typical non-rigid variations in cochlear anatomy. Once it is non-rigidly fitted to the cochlea shape in a new patient’s CT, it allows accurately estimating the position of fine-scale internal cochlear structures that are not directly visible in the patient’s CT. The model fitting of the cochlear structures was based on pre-implantation images when available. When no pre-implantation CT was available, either the mirror image of the opposite unimplanted cochlea (for SSD-CI subjects; Reda et al., 2014) or a machine-learning-based approach using the post-implantation CT alone (for BI-CI subjects; Wang et al., 2019) was used. The electrode array was localized in a post-implantation CT using automated algorithms (Zhao et al., 2019). The two results were then merged using well-known rigid image-registration techniques (Maes et al., 1997) as has been validated in histological studies (Schuman et al., 2010).

The modeling analysis quantifies the estimated electrode position in three dimensions (Verbist et al., 2010): (1) the insertion angle, based on a coordinate system defined by a line drawn between the round window and the modiolus, (2) the distance between the electrode and the modiolus (i.e., the modiolar distance), and (3) the scalar location of the electrode (i.e., whether it is located within scala tympani, within scala vestibuli, or in the in-between region designated as “scala media or basilar membrane”). In this study, the main outcome measure of interest for the purposes of comparing estimates of interaural place mismatch was the insertion angle. For the BI-CI subjects, the matching comparison electrode for a given reference electrode was extracted by linearly interpolating the insertion angle data, then finding the comparison electrode at the same insertion angle as the reference electrode. For the SSD-CI subjects, the matching acoustic frequency for a given reference electrode was derived using the Stakhovskaya et al. (2007) spiral-ganglion correction to the Greenwood map.

### Experimental Design and Statistical Analysis

For each reference electrode tested, an estimate of the matched comparison electrode or comparison acoustic frequency was extracted for each of the three methods (ITD, CT, and Pitch) as described above. Only those electrodes for which matching estimates were available for all three measures were included in further data analysis (because of its time-consuming nature, fewer electrodes were tested in the ITD experiment). For each measure, the match in the comparison ear was converted into an equivalent insertion angle as described below in the “*Relationship between the three measures”* subsection of the Results. Using these estimates, two sets of analyses were carried out using linear mixed-model regression *(lmer* function in Rstudio version 1.2.5001).

#### Across-subject relationship between measures

The purpose of this analysis was to determine the extent to which each measure yielded a similar estimate of mismatch. The magnitudes of the interaural place mismatch-estimates were compared in three mixed-model analyses, each making pairwise comparisons between two of the mismatch measures across the electrodes. Each model treated one mismatch estimate as the outcome variable, the other as a fixed effect, group (BI-CI or SSD-CI) as another fixed effect, and subject as a random effect. Bonferroni corrections were applied for three comparisons (criterion: p<0.05/3=0.0167). Main effects and interactions are reported in terms of chi-squared values describing the effect of the removing the term of interest from the model.

#### Tonotopic dependence

The purpose of the analysis was to determine how the three estimates of mismatch vary as a function of tonotopic location along the electrode array. An initial linear mixed-model regression analysis was carried out to examine the dependence of mismatch on the interaural matching measure (ITD, CT, or Pitch), with tonotopic location (defined by the clinical CF associated with each reference electrode), and subject group (SSD-CI or BI-CI) as fixed effects, and subject included as a random effect. Following up on observed significant interactions, separate Bonferroni-corrected analyses (criterion: p<0.05/2=0.025) were carried out for each subject group, which were the same as for the initial analysis except that subject group was not included as a factor.

## Results

### Example individual results

The study results are organized by first showing example findings for each of the three measures of interaural place mismatch for three sample BI-CI and three sample SSD-CI subjects (Fig. 1), with each column showing the data for one subject. These six individuals were chosen to highlight the range of outcomes revealed by the three measures. In this figure and throughout the results, the horizontal axis is arranged such that the lowest-frequency electrode (or lowest acoustic frequency) toward the apex of the cochlea is on the left. For Cochlear-brand devices, the electrodes are numbered in descending order from 22 (low) to 1 (high frequency), while for MED-EL devices, the electrodes are numbered in ascending order from 1 (low) to 12 (high frequency). Figure 1A-B shows example results for ITD discrimination; Figure 1C-D shows example results for pitch ranking; Figure 1E-F shows example CT scan results. The bottom row (Fig. 1G-H) plots all three estimates of the relative place of electrical stimulation as estimated by the three different measures.

**Figure 1.**
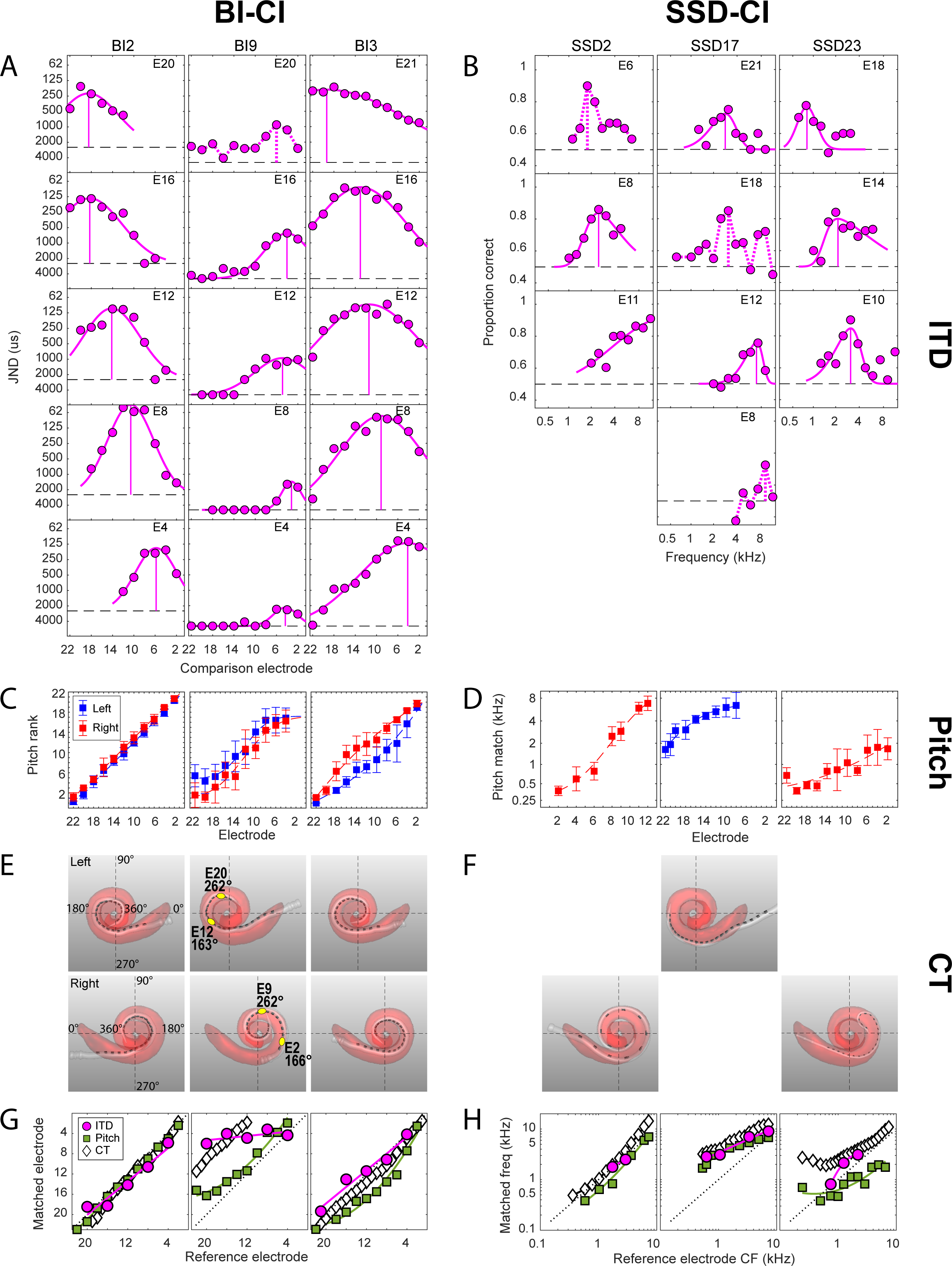
Examples of the perceptual and objective data collected for the three estimates of interaural place mismatch for six example subjects (three BI-CI and three SSD-CI). ***A- B*,** ITD discrimination data: JND (BI-CI subjects) or proportion correct as a function of the comparison electrode or frequency for a given reference electrode. ***C-D,*** pitch ranking data: rank (BI-CI subjects) or pitch match (SSD-CI subjects) as a function of electrode number. ***E-F*,** computational model output of the CT scan analysis, showing the estimated cochlear morphology (red), and the position of the electrode array (white) and individual electrode contacts (dark gray). ***G-H*,** summary of the three estimates of interaural place match as a function of reference electrode number (BI-CI subjects) or clinical CF (SSD- CI subjects) derived from the measurements in ***A-F***. The diagonal dotted lines in ***G-H*** represent perfect interaural alignment; vertical displacement from this line represents mismatch.

#### Interaural time-difference discrimination

Figure 1A-B shows example ITD- discrimination tuning curves, with each panel representing a difference reference electrode, arranged from top to bottom from apex (low-frequency electrode) to base (high-frequency electrode). For the BI-CI subjects, the individual panels in Fig. 1A indicate the ITD JND for left/right discrimination as a function of the comparison electrode number. For the SSD-CI subjects, the individual panels in Fig. 1B indicate the proportion-correct performance (in identifying the interval that contained an ITD that changed across the four bursts) as a function of the CF of the acoustic click train presented to the unimplanted ear.

The key parameter that was extracted from the fitted functions for further analysis was the comparison electrode or acoustic frequency that yielded peak performance, which is identified by a vertical line in each panel of Fig. 1A-B. In some cases (6% of electrodes tested), there was a prominent peak in the performance function, but a skewed-Gaussian curve could not be fit to the function. In these cases, the acoustic comparison frequency yielding a peak in the performance function was taken as the frequency-match estimate. These cases are identified by dashed magenta lines in Fig. 2A-B. There were also several cases (13% of electrodes tested) where no frequency-match estimate was made (Bernstein et al., 2018), either because the performance function showed no prominent peak, showed two or more peaks of similar amplitude, or suggested a peak outside of the comparison range. In these cases, identified by the absence of a vertical magenta line in Fig. 2A-B, the electrode was discarded from further analysis.

**Figure 2.**
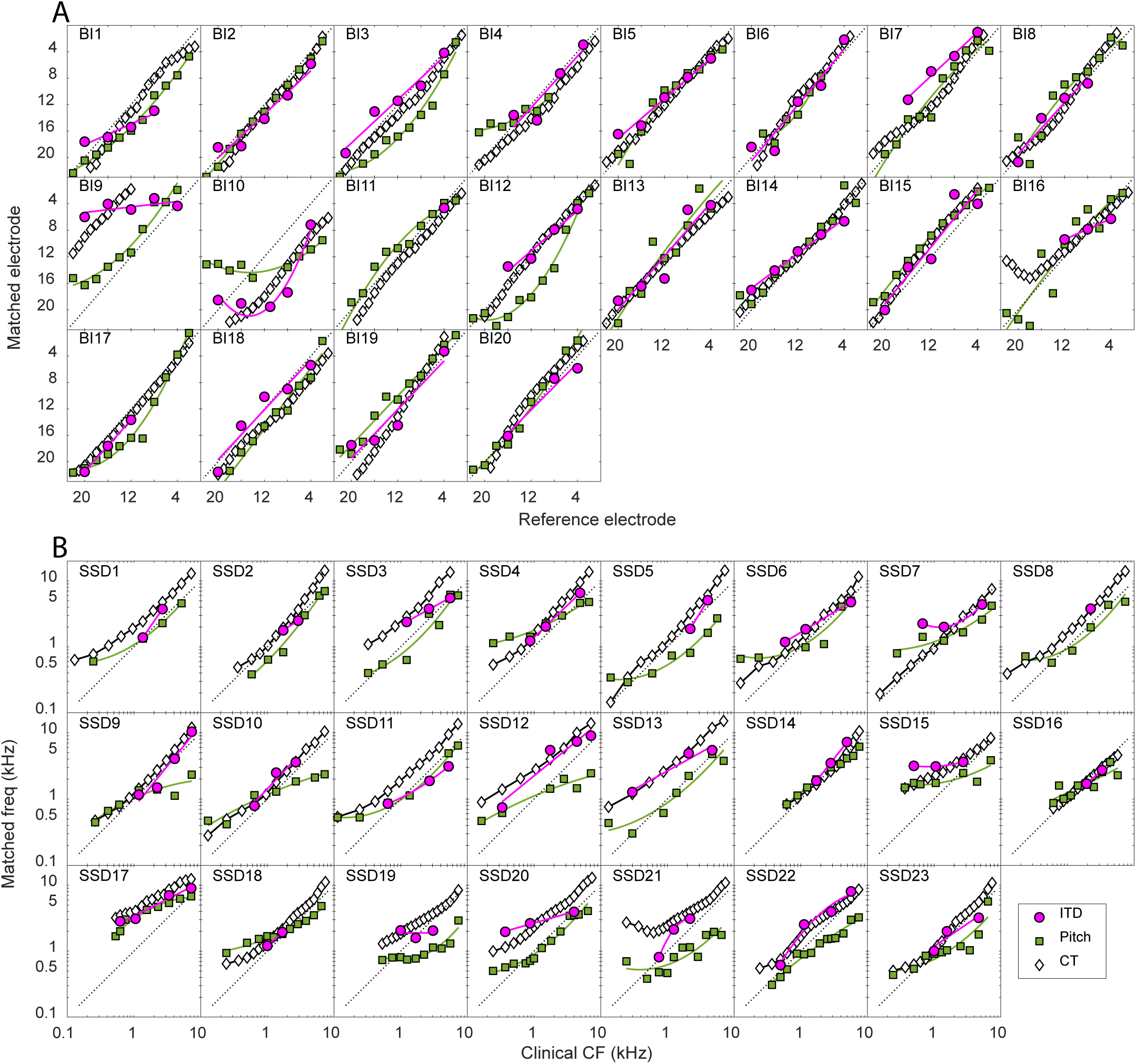
Summary of the three estimates of interaural place for each individual listener in the study. ***A*,** For the BI-CI listeners, the matched electrode in the comparison ear is plotted as a function of the reference electrode. ***B***, For the SSD-CI listeners, the matched acoustic frequency in the comparison ear is plotted as a function of the clinical CF associated with the reference electrode. The diagonal dotted lines represent perfect interaural alignment; vertical displacement from this line represents mismatch.

#### Pitch ranking

Figure 1C-D shows example results for the pitch-ranking experiment. The BI-CI subjects jointly ranked all even electrodes in both ears. Therefore, there are two curves indicating the average rank for the electrodes in each ear in Fig. 1C. For the BI-CI subjects, the pitch match for each electrode in the reference ear was derived by fitting sigmoidal curves the data for each ear, and then extracting from these fitted functions the comparison electrode that yielded the same pitch rank as the reference electrode in question. Figure 1C shows a variety of outcomes for the pitch-matching task, from a case with very little mismatch (BI2, left column) to cases with substantial mismatch that either varied (BI3, right column) or was reasonably constant (BI9, middle column) across the array.

The SSD-CI subjects performed the pitch-ranking task for each reference electrode relative to a set of frequencies that was assumed to be correctly ordered in the acoustic ear. Therefore, there is only one pitch ranking curve for each subject in Fig. 1D. This figure shows that there was a wide variety of pitch-match estimates across the three example SSD-CI subjects. For subject SSD2 (left panel), the pitch matches across the array covered nearly the full range of comparison frequencies. For the other two subjects, the range of matches was limited to the upper (SSD17, middle panel) or lower (SSD23, right panel) portions of the comparison range.

#### Computed-tomography scans

A visual depiction of the CT scan modeling analysis is shown for both ears for the three example BI-CI subjects (Fig. 1E) and for the implanted ear for the three example SSD-CI subjects (Fig. 1F). These images include the estimated cochlear morphology (pink shading), the estimated location of the array (white) and individual electrodes (dark gray) within the cochlea. These examples show a variety of insertion depths and mismatch.

For the BI-CI subjects (Fig. 1E), there was variety both in the absolute insertion depth and the degree of mismatch in the insertion between the two ears. The left (BI2) and right (BI3) columns show an insertion of approximately one full turn that was similar between the two ears. The middle column (subject BI9) shows a deep insertion of more than a full turn in the right ear, but a shallower insertion of about a full turn in the left ear. As a result, this subject has 10-11 electrodes of interaural place mismatch (yellow points in Fig. 1E; black diamonds in Fig. 1G). Reference (left) E12 and comparison (right) E2 had matching insertion angles (163-166°), as did reference E20 and comparison E9 (262°). For the SSD-CI subjects, Fig. 1F shows a relatively deep insertion greater than one full turn (SSD2, left column), a relatively shallow insertion of only about half a turn (SSD17, middle column), and a case of limited insertion depth due to a tip foldover (SSD23, right column).

#### Estimates of interaural place mismatch

Figure 1G-H plots the estimates of the relative place of electrical stimulation as estimated by the three different measures. For the BI-CI subjects, the horizontal axis represents the reference electrode, while the vertical axis represents the matching comparison electrode in the other ear. For the SSD-CI subjects, the horizontal axis represents the reference-electrode CF based on the subject’s clinical map, while the vertical axis represents the estimated matched acoustic frequency plotted on the Greenwood scale. The diagonal dashed line represents the 1:1 line of equality; in other words, where the comparison electrode or acoustic frequency was matched to the reference electrode or acoustic frequency. Points falling above the diagonal line indicate cases where a reference electrode was stimulating more basally (i.e., a higher frequency cochlear place) than the same electrode number or acoustic frequency presented to the comparison ear. For the SSD-CI subjects, mismatch is expected to be in the basal direction, as the array is generally not inserted deeply enough to stimulate the apical portion of the cochlear (Landsberger et al., 2015). For the BI-CI subjects, a mismatch could go in either direction, depending on the relative insertions in the two ears.

For the example subjects depicted in the left columns for each subject group in Fig. 1 (BI2 and SSD2), all three estimates were relatively consistent with one another and there was little interaural place mismatch. For the example subjects depicted in the center columns for each subject group (BI9 and SSD17), all three measures showed substantial interaural place mismatch. For BI9, the three estimates were not closely aligned even though all three suggested a mismatch. For SSD17, the three measures showed similar estimates of electrode position. For the example subjects depicted in the right columns for each subject group (BI3 and SSD23), the three measures suggested different mismatch results. For BI3, the CT scans suggested little or no mismatch, while the ITD and Pitch measures suggested that there was some mismatch, but in opposite directions. For SSD23, the CT scan suggested substantial mismatch, the Pitch measure suggested mismatch in the opposite direction, and the ITD measure suggested a mismatch that was similar to the CT scan for two of the electrodes tested, but no mismatch for the third electrode.

### Relationship between the three measures

#### Individual data

All three estimates of the interaural place-of-stimulation mismatch are plotted for each individual subject in Fig. 2. These plots are similar to the summary plots and follow the same plotting conventions as for the example subjects in Fig. 1G-H, except here, the individual results are shown for all 20 BI-CI and 23 SSD-CI subjects in the study.

Several trends are apparent in these individual data. First, the BI-CI subjects (Fig. 2A) showed relatively little mismatch overall, while many of the SSD-CI subjects showed substantial mismatch (Fig. 2B). This was especially the case for the CT (black diamonds) and ITD measures (magenta circles). For the BI-CI subjects, the vast majority of the place estimates fell within ±2 electrodes of the diagonal (with some exceptions, for example, subjects BI9 and BI10), whereas for the SSD-CI subjects, the vast majority of the estimates fell above the diagonal line. Second, the CT and ITD estimates were generally consistent with one another. For the BI-CI subjects, they both fell near the diagonal in most cases, indicating little interaural place mismatch, whereas for the SSD-CI subjects, these two estimates tended to have a similar slope and show a similar degree of excursion from the diagonal. There were, however, exceptions. For example, the CT scan suggested a large mismatch but the ITD measure did not for subject SSD11. Third, the Pitch estimates (green squares) were much more variable than and were often inconsistent with the other two estimates. For the BI-CI subjects, there were a number of cases where the CT and ITD measures suggested little or no mismatch, while the Pitch measure suggested substantial mismatch (e.g., subjects BI3 and BI12). For the SSD-CI subjects, the Pitch estimates were in general lower than the other two estimates, and with a more gradual slope, suggesting positive mismatch for more apical electrodes, and negative mismatch for basal electrodes.

#### Group data

To compare the three measures, the CT-based (dCT), ITD-based (dITD), and Pitch-based (dPitch) estimates of interaural place mismatch were defined for a given reference electrode as the distance between the cochlear locations associated with each estimate and the clinical map. All mismatch estimates are expressed in terms of insertion angle. For the BI-CI subjects, the comparison-ear CT scan formed the basis of the translation between electrode number and insertion angle. Mismatch was calculated by comparing the insertion angle of the matched comparison electrode to the insertion angle of comparison electrode with the same number as the reference. For the SSD-CI subjects, the Stakhovskaya et al. (2007) spiral-ganglion map formed the basis of the translation between matched acoustic frequency and insertion angle. Mismatch was calculated by comparing the insertion angle associated with the matched acoustic frequency to the insertion angle associated with reference electrode clinical CF. For both groups, the sign convention was established such that a positive value always reflected the expected direction of the mismatch. For the BI-CI subjects, the expected direction for a given subject was determined from the CT scan by estimating the average mismatch across the array. For the SSD-CI subjects, the expected direction was always defined relative to the clinical CF, based on the assumption that the cochlear place of electrical stimulation would generally be basal (i.e., higher frequency) to the CF-equivalent place (Landsberger et al., 2015).

The pairwise relationships between the three measures of mismatch are plotted in Fig. 3. The BI-CI results are shown in the top row (Fig. 3A-C) and the SSD-CI results are shown in the middle row (Fig. 3D-F), with each symbol/color combination in a given row representing a different subject and with each data point representing one individual electrode. The results for the two groups are combined in the bottom row (Fig. 3G-I). Data are plotted for only those reference electrodes where data were available for all three measures. The shaded squares near the center of Fig. 3A-F represents the “binaural tolerance” mismatch range of ±75°, which is roughly equivalent to the ±3 mm on the Greenwood scale over which binaural sensitivity has previous been shown to be insensitive to mismatch (Kan et al., 2013). Points falling within this square indicate electrodes for which both measures show an interaural place mismatch that is smaller than this tolerance range. The thick diagonal line in each panel represents a 1:1 correspondence (i.e., the two measures in question give the same estimate of interaural mismatch), while the two thinner diagonal lines indicate the ±75° range around the center diagonal.

**Figure 3.**
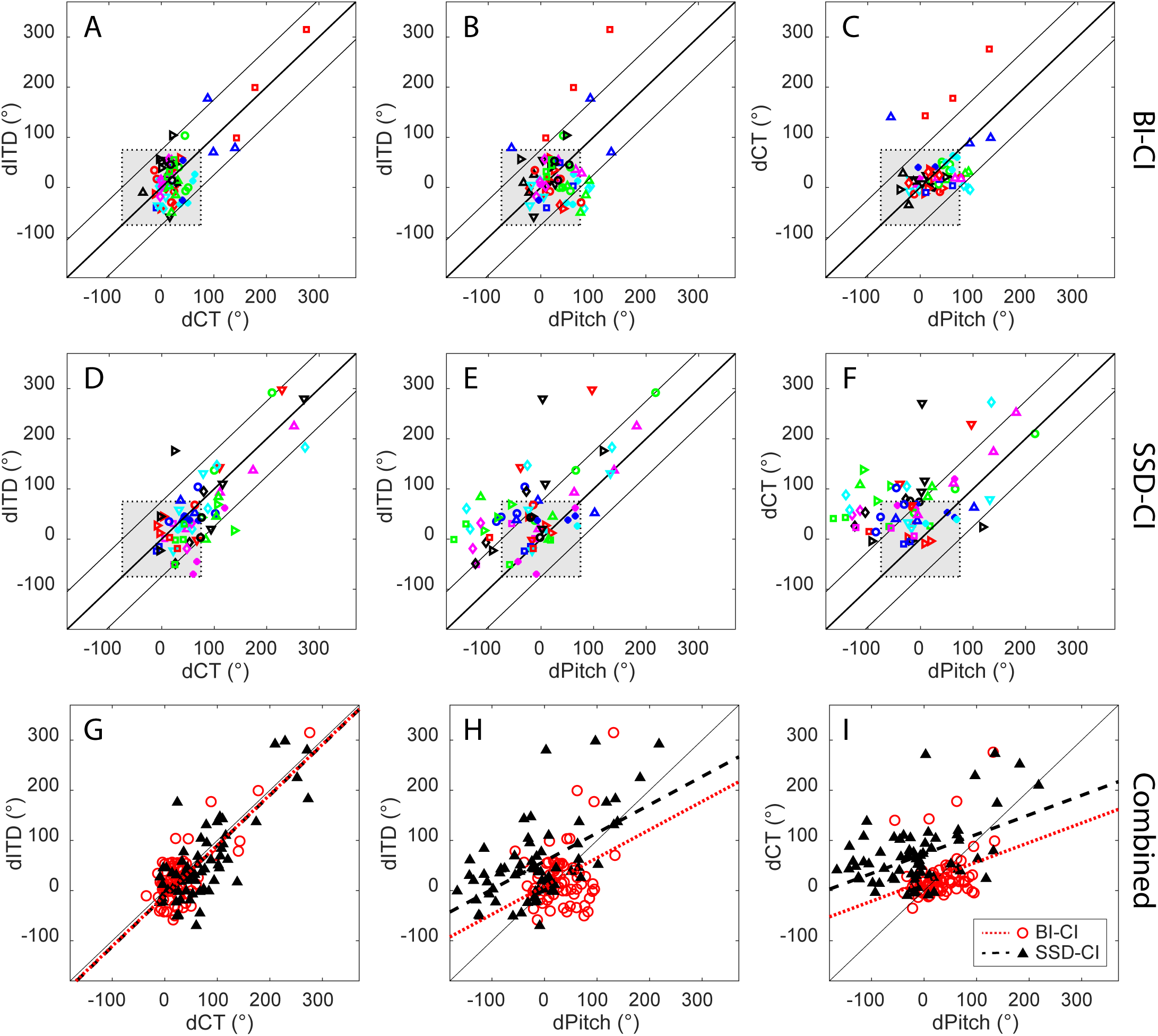
Pairwise comparisons of the three estimates of the magnitude of interaural place mismatch for individual electrodes for: ***A-C*,** the BI-CI subjects, with each color/symbol combination representing a different subject; ***D-F***, SSD-CI subjects, with each color/symbol combination representing a different subject and ***G-I***, both groups combined. The diagonal lines indicate 1:1 correspondence (±75°) where two mismatch estimates are equal. The grey shaded box indicates the ±75° mismatch range (equivalent to 3 mm in the Greenwood scale) over which binaural sensitivity is estimated to be tolerant to mismatch (Kan et al., 2013).

For the BI-CI subjects (Fig. 3A-C), the estimated interaural mismatch was outside the 75° tolerance range for fewer than 1/6 of the electrodes tested (ITD: 11%; CT: 8%; Pitch: 15%). For the SSD-CI subjects (Fig. 3D-F), mismatch estimates fell outside of the 75° tolerance range for 1/4 to 1/3 of the electrodes tested (ITD: 25%; CT: 36%; Pitch: 36%). Comprehensive analyses are described below that examine the tonotopic relationships between the three measures. It was nevertheless instructive to carry out an initial set of pairwise analyses get a sense of the relationships between the three measures of mismatch. Bonferroni-corrected linear mixed-model regression analyses were carried out for each of the three pairwise comparisons of mismatch, taking into account the data from both subject groups (Fig. 3G-I). In each analysis, the estimate on the vertical axis was treated as the outcome variable, the estimate on the horizontal axis as a fixed effect, group as another fixed effect, and subject as a random effect. To provide a sense of the strength of the pairwise relationships, estimated marginal R^2^ values associated with adding the variable represented on the horizontal axis in the model are reported (as derived using the *r2* function from the *sjstats* library for R) in addition to the fixed-effects statistics.

The left column of Fig. 3 shows that dITD and dCT were closely related for both groups. There was no significant main effect of group [χ^2^(1)=2.14, p=0.14] or interaction between group and dCT [χ^2^(1)=0.18, p=0.67], indicating that the relationship between dITD and dCT was the similar for the two groups. There was a strong, significant relationship between dITD and dCT [Fig. 3G; t(101)=13.7, p<0.0001; R²=0.63] and these two mismatch estimates were nearly equal, with a regression slope that was not significantly different from unity (B=0.96, p=0.58). This relationship is seen clearly in Fig. 3G, where the points for both subject groups are near the 1:1 line.

The middle and right columns of Fig. 3 show the relationships between dPitch and the other two measures. While there was no significant interaction between group and dPitch in either case [dCT vs. dPitch: χ^2^(1)=2.30, p=0.13; dITD vs. dPitch: χ^2^(1)=1.26, p=0.26], there were significant main effects of group [dCT vs. dPitch: χ^2^(1)=13.7, p=0.002; dITD vs. dPitch: χ^2^(1)=10.8, p=0.001]. This reflects the observation that for a given amount of mismatch as estimated by ITD (Fig. 3H) or CT (Fig. 3I), the SSD-CI subjects (black triangles) showed less (or more negative) mismatch as estimated by Pitch than the BI-CI subjects (red circles). There was a significant relationship between dPitch and dCT [Fig. 3H, χ^2^(1)=42.0, p<0.0001, R^2^=0.27] and between dPitch and dITD [Fig. 3I, χ^2^(1)=43.2, p<0.0001, R^2^=0.28]. Despite these significant relationships, many of the points fell outside the region indicated by the upper and lower diagonal lines in each panel, especially for the SSD-CI subjects (Fig. 3E-F), indicating the that dPitch differed from dITD or dCT by more than 75°. Furthermore, the regression slopes involving dPitch were significantly smaller than unity (dPitch vs. dCT: B=0.39; dPitch vs. dITD: B=0.56; p<0.0001 in both cases). Thus, considering the scatter in the data and the shallow slopes for each relationship, dPitch differed substantially from these other estimates despite the significant correlations.

### Tonotopic dependence of interaural frequency mismatch

Figure 3 does not account for how the relationship between mismatch estimates might change as a function of intracochlear location. For example, for many SSD-CI subjects (Fig. 2B), dPitch (green squares) had a shallower slope with respect to the clinical CF than the slope of dCT (black diamonds) or the slope of dITD (magenta circles). This suggests that the relationship between the mismatch estimates was dependent on the cochlear region being stimulated.

The three estimates of interaural frequency mismatch are plotted as a function of location along CI array for the BI-CI subjects (Fig. 4A-E) and for the SSD-CI subjects (Fig. 4F-J). A main question posed in this study was whether the Pitch and ITD measures of electrode position aligned more closely with the clinical CF (indicating that the percept had shifted to align with any existing mismatch) or more closely with the CT-scan estimate (indicating a lack of plasticity to any existing frequency mismatch). To pose this question, mismatch was defined in two different ways. The first (Fig. 4A-C) and third rows (Fig. 4F-H) plot the estimated mismatch with respect to the reference CI electrode (electrode number for the BI-CI subjects; clinical CF for the SSD-CI subjects). Here, mismatch estimates near zero indicate close alignment with the clinical CF. The second (Fig. 4D-E) and fourth rows (Fig. 4I-J) plot the estimated mismatch with respect to the insertion angle estimates from the CT scan. Here, mismatch estimates near zero indicate close alignment with physical electrode location. The three curves in each panel of Fig. 4 represent the estimated group mean and 95% confidence interval of the mismatch for a given cochlear location. Electrode locations for which the 95% confidence interval does not include zero indicate a significant group-average mismatch.

**Figure 4.**
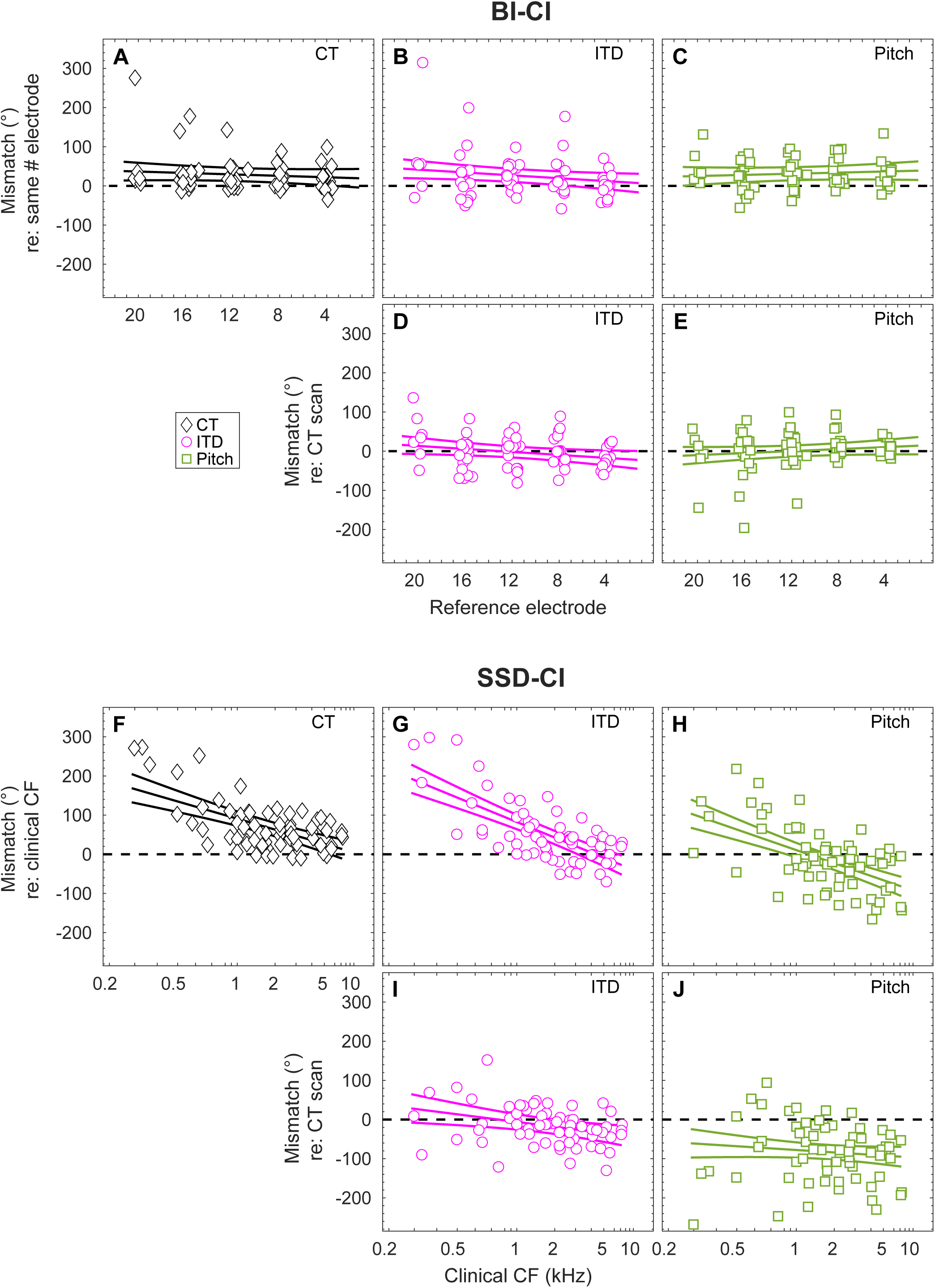
Tonotopic dependence of interaural place mismatch for ***A-E*,** BI-CI subjects and ***F-J*,** SSD-CI subjects. The first and third rows reference the mismatch to the clinical map, based on: ***A-C*** the insertion angle associated with the number-matched electrode for BI- CI subjects or ***G-H*** the insertion angle associated with the clinical CF for SSD-CI subjects. The second and fourth rows (***D-E,I-J*)** reference the mismatch to the CT estimate of electrode position. Points represent mismatch measurements for individual electrodes; fitted curves represent estimates of the group-average and 95% confidence interval.

An initial linear mixed-model regression analysis examined the dependence of mismatch on the interaural matching measure, tonotopic location (clinical CF for each reference electrode), and subject group. There was no significant three-way interaction [χ^2^(2)=4.65, p=0.10] or two-way interaction between tonotopic location and measure [χ^2^(2)=1.18, p=0.56]. However, there were significant two-way interactions between tonotopic location and group [χ^2^(1)=101, p<0.0001] and between measure and group [χ^2^(2)=42.8, p<0.0001], confirming that the dependence of mismatch on measure and tonotopy was different for the two groups. Therefore, separate Bonferroni-corrected analyses were carried out for the two subject groups.

For the BI-CI subjects, all three estimates of interaural mismatch gave similar average results (Fig. 4A-C). A linear mixed-model regression analyses with two fixed-effect factors (interaural matching measure and electrode number) found no significant main effect of electrode number [χ^2^(1)=1.59, p=0.21], main effect of interaural matching measure [χ^2^(2)=1.01, p=0.60], or interaction between these two factors [χ^2^(2)=4.73, p=0.094]. The y-intercept was significant [χ^2^(1)=9.94, p=0.0016] with a value of 28.6°. While this suggests that there was, on average, some interaural frequency mismatch for the BI-CI subjects, this significant difference is not necessarily meaningful because the mismatch direction was always defined to be positive for the CT-scan electrode-position estimates for each subject. When calculated relative to the CT estimates of electrode position (Fig. 4D-E), neither dPitch nor dITD was significantly different from zero. In summary, this means that on average across the subjects, none of the three measures of mismatch were different from one another.

For the SSD-CI subjects, the mismatch estimates clearly differed between measures (Fig. 4F-H). This observation was supported by a linear mixed-model regression analyses carried out on the mismatch estimates with respect to each electrode’s clinical CF. There were significant main effects of CF [χ^2^(1)=112, p<0.0001] and measure [χ^2^(2)=54.7, p<0.0001], but no significant interaction between the two variables [χ^2^(2)=3.45, p=0.18].

For all three measures, the amount of mismatch was more positive at the apical end than the basal end of the array. Relative to the clinical CF, dCT was significantly greater than zero across the length of the array (Fig. 4F), indicating that the electrode location was consistently basal to the clinical CF as expected, although the mismatch was small at the basal end. Likewise, dITD (Fig. 4G) was significantly greater than zero across the apical and middle portions of the array, becoming non-significant at the basal end. In contrast, the direction of dPitch varied across the electrode array (Fig. 4H). At low frequencies, there was mismatch in the expected positive (basal) direction, while at high frequencies there was mismatch in the unexpected negative (apical) direction.

Relative to the CT scan, dPitch (Fig. 4J) was significantly negative for the full frequency range, meaning that the pitch associated with a given electrode was lower than expected given the electrode position. In contrast, dITD (Fig. 4I) was not significantly different than zero for most of the array—meaning that binaural processing was optimal for an acoustic frequency at the electrode position indicated by the CT scan—except at the very basal end of the array, where dITD was slightly negative.

In summary, there was on average very little mismatch for the BI-CI subjects, and no discernable difference between dITD, dPitch and dCT (Fig. 4A-C). In contrast, all three measures showed considerable group-average mismatch for the SSD-CI subjects that varied across the length of the electrode array (Fig. 4F-H). The Pitch estimates aligned with neither the clinical CF (Fig. 4H) nor the CT-estimated electrode position (Fig. 4J). The ITD and CT estimates were closely aligned: relative to the clinical CF (Fig. 4F-G), both measures showed a large average mismatch at the apical end of the array that decreased to near zero at the basal end. When referenced to the CT estimate of electrode position, the ITD measure showed no significant mismatch except for a small negative mismatch at the basal end (Fig. 4I).

## Discussion

This study asked if the binaural system and place-pitch perception are sufficiently plastic to adapt to interaural place mismatch. All subjects had >6 months of experience using their CIs, with most subjects having >1 year, whereas complete adaptation, at least for CI speech understanding, is thought to occur for post-lingually deafened adults by 6 months for BI-CIs (Reeder et al., 2014) and 3 months for SSD-CIs (Buss et al., 2018). Plasticity over this period of device use can be inferred from comparisons between the mismatch measures: a psychoacoustic estimate that aligns with the CT estimate of mismatch could be interpreted as a lack of plasticity to mismatch; a shift toward the clinical CF could indicate adaptation has occurred (Reiss et al., 2014).

CT and ITD estimates of interaural place mismatch were closely aligned (Fig. 3G), suggesting a lack of binaural-system plasticity to mismatch. For the BI-CI subjects, although few of the electrodes tested showed substantial mismatch, these two mismatch estimates were consistent for this subset of cases (Fig. 3A). While the relative lack of mismatch made it difficult to assess tonotopic dependence, the CT and ITD estimates showed a similar (small) magnitude of group-average mismatch for number-matched electrodes (Fig. 4A,B,D).

For the SSD-CI subjects, many electrodes showed mismatch >75° and the ITD and CT scans gave similar estimates (Fig. 3D). Furthermore, both measures showed similar tonotopic dependence, with mismatch largest toward the apex and decreasing to zero toward the base (Fig. 4F-G). This tonotopic dependence differs from other radiographic studies of acoustic-electric mismatch for monaural CI users that have found little tonotopic dependence (Landsberger et al., 2015; Canfarotta et al., 2020), except for a slight tendency for more mismatch near the apex. Note that these previous studies based their estimates on the manufacturer’s standard frequency allocation. The current study was specifically interested in comparing mismatch estimates relative to the actual clinical map and several SSD-CI subjects had one or more basal electrodes deactivated clinically due to incomplete insertion, high impedance, or perceptual annoyance. As a result, CFs for the most basal active electrodes were shifted upward from the manufacturer default, likely explaining some of the discrepancy.

By comparing binaural tuning to radiographic images of physical electrode position, these results extend previous findings of frequency-tuned binaural sensitivity for BI-CI (Hu and Dietz, 2015; Kan et al., 2015), SSD-CI (Bernstein et al., 2018; Francart et al., 2018; Dirks et al., 2020), and bimodal-CI subjects (Francart et al. 2011, 2014). Hu and Dietz (2015) found that electrophysiological and psychophysical estimates of mismatch for binaural processing were consistent with each other, but differed from pitch-based estimates that showed little mismatch, suggesting a lack of BI-CI binaural plasticity to mismatch. However, the small sample (N=7, one electrode per subject) precluded statistical assessment of these relationships and, without imaging data regarding electrode positions, these data provided only indirect evidence of a lack of binaural plasticity.

While the auditory system exhibits central plasticity following peripheral changes stemming from hearing loss and CIs (Fallon et al., 2009), it is unclear if there is sufficient plasticity to overcome interaural place mismatch in adult humans (Kan et al. 2013, 2015; Reiss et al. 2014, 2015; Aronoff et al. 2016, 2019). Other dimensions of interaural mismatch might be more readily rectified by plasticity (e.g., King *et al*., 2001). For example, adaptation to altered ITDs or interaural level differences appears to occur behaviorally in normal-hearing human adults (Shinn-Cunningham et al. 1998a,b; Keating et al. 2016) and juvenile ferrets (Keating et al., 2013), and neurophysiologically through altered spatial maps in the optic tectum for developing owls (Mogdans and Knudsen, 1992; Linkenhoker and Knudsen, 2002) and in the juvenile and adult ferret cortex (Keating et al., 2013, 2016). However, there is scant evidence for plasticity to interaural place mismatch. Our data suggest that plasticity does not overcome interaural place mismatch for binaural tuning (Figs. 2-4).

In contrast to the close relationship between the ITD and CT measures, there was little correspondence between Pitch and these other measures. While the Pitch estimates were significantly correlated with ITD and CT estimates, the slopes were much less than one (Fig. 3H-I). Furthermore, Pitch mismatch was often negative (Fig. 3B-C,E-F,H-I), opposite from the expected direction (for BI-CI subjects, a positive mismatch is expected because the direction is referenced to the CT-scan mismatch direction; for SSD-CI subjects, a positive mismatch is expected because the array does not reach the apex of the cochlea).

For the BI-CI subjects, there was a small amount of average Pitch mismatch when referenced to the matched electrode number (Fig. 4C), but none when referenced to the CT scan location, suggesting no plasticity to any small mismatch that was present. For the SSD-CI subjects, there was considerable Pitch mismatch when referenced to the clinical CF (Fig. 4J), yet the average mismatch did not align with the CT (Fig. 4H). While the lack of correspondence with CT estimates might suggest plasticity, the Pitch estimates were consistently lower than even the clinical CF in the basal half of the array (Fig. 4H), which is inconsistent with a hypothesized shift in place pitch toward the clinical CF.

Overall, the literature is mixed regarding the question of pitch plasticity following CI use. Some studies have shown plasticity toward clinical CFs (Reiss et al., 2014; Hu and Dietz, 2015; Tan et al., 2017) while others have shown little such evidence. For example, Aronoff et al. (2016) found that BI-CI pitch matches often deviate from number-matched electrodes, while Schatzer et al. (2014) and Marozeau et al. (2020) found that SSD-CI pitch matches align more closely with radiographic estimates of electrode position than with clinical CFs.

One possible reason for the disparate results across studies is that the procedures (Jensen et al., 2021) and stimuli (Adel et al., 2019) employed had a large impact on the observed results, which questions whether these measurements reflect place of stimulation. In the current study, the surprising tonotopic pattern of pitch-match estimates for SSD-CI listeners (positive at the apex, negative at the base, Fig. 4H) might reflect a procedural bias where the pitch match gravitated toward the center of the range of available comparison frequencies (Carlyon et al., 2010; Goupell et al., 2019).

Despite the clear relationship observed between ITD and CT-scan estimates of mismatch, but not Pitch, there were at least four study limitations that provide clear future directions. First, this study was performed acutely. There was a range of CI experience (Tables I and II), with many subjects having used their CI(s) for multiple years, but some for as little as 6 months. It is possible that any adaptation these subjects were experiencing was incomplete, particularly for binaural processing where BI-CI functional performance can continue to improve for up to 4 years (Eapen *et al*., 2009). Any clinical changes to CI sound-processor maps might also affect this timeline. A longitudinal study of changes in ITD and pitch over the course of years could address this limitation.

Second, it is unclear if the pitch results reflect plasticity or procedural biases, which are common and often large for CI subjects (Carlyon et al., 2010; Goupell et al., 2019; Jensen et al., 2021). Nearly all the individual pitch-match estimates here passed the bias check for independence from the adaptive-track starting point (Carlyon et al., 2010). Yet Jensen et al. (2021) found that adaptive measures can pass this check and still be susceptible to other biases. While Jensen et al. found the ranking procedure employed here to be largely immune to systematic biases for BI-CI subjects, that study did not include SSD-CI subjects. Further work is required to evaluate possible biases for SSD-CI subjects. Confirming pitch plasticity in a longitudinal study could help to clarify this issue.

Third, this study did not estimate possible local degeneration of the spiral ganglion population (Long *et al*., 2014). CT images identify electrode placement, but do not assess the number and health of the spiral ganglia, the cells being electrically excited. An assessment of neural survival, for example using electrically evoked compound action-potential measurements, might account for some of the deviation between the CT and perceptual mismatch measures (Bierer, 2010; Long et al., 2014; DeVries et al., 2016).

Fourth, most of the BI-CI subjects in this study had little appreciable interaural place mismatch (Fig. 2-4). Future work examining BI-CI subjects with larger interaural place mismatch would strengthen any claims concerning lack of binaural plasticity. In closing, it is important to note that this study serves a practical purpose by informing BI-CI and SSD-CI clinical practice. The finding that binaural sensitivity appears to not adapt to interaural place mismatch means that maximizing binaural performance will likely require intervention by adjusting clinical CFs. CT-scan estimates of interaural place mismatch showed relatively close agreement with time-consuming ITD-based estimates (Figs. 3-4). This suggests that CT imaging may prove to be an effective clinical tool to measure interaural place mismatch, guiding the audiologist in frequency mapping to optimize binaural processing without requiring extensive psychophysical testing.

## Data Availability

The raw data were generated at Walter Reed National Military Center and the University of Maryland - College Park. The authors confirm that the derived data supporting the findings of this study are available within the article.

## ACKNOWLEDGMENTS

Research reported in this publication was supported by the National Institute On Deafness And Other Communication Disorders of the National Institutes of Health under Award Numbers R01 DC015798 (J.G.W.B. and M.J.G.) and R01 DC014037 (J.H.N.). The content is solely the responsibility of the authors and does not necessarily represent the official views of the National Institutes of Health.

We thank Cochlear Ltd. and Med-El for testing equipment and technical support. We thank Coral Dirks, Ginny Alexander, Kelly Johnson, Danielle King, Taylor Bakal, and Stefano Cosentino who helped to collect and analyze data for this study, and Elicia Pillion, Natalia Stupak, John Galvin, René Gifford, Elizabeth Searing, and Sarah Natale for assistance with subject recruitment. We thank Ginny Alexander for managerial help.

This work was partially presented at the 175th Meeting of the Acoustical Society of America, Minneapolis, MN (May 2018), the Association for Research in Otolaryngology 42nd MidWinter Meeting, Baltimore, MD (February 2019), and the Conference on Implantable Auditory Prostheses in Lake Tahoe, CA, USA (July 2019).

The identification of specific products or scientific instrumentation is considered an integral part of the scientific endeavor and does not constitute endorsement or implied endorsement on the part of the authors, Department of Defense, or any component agency. The views expressed in this article are those of the authors and do not reflect the official policy of the Department of Army/Navy/Air Force, Department of Defense, or U.S. Government.

## Notes

### Competing Interest Statement

The authors have declared no competing interest.

### Funding Statement

Research reported in this article was supported by the National Institute On Deafness And Other Communication Disorders of the National Institutes of Health under Award Numbers R01 DC015798 (J.G.W.B. and M.J.G.) and R01 DC014037 (J.H.N.).

### Author Declarations

Research was approved by the IRBs for Walter Reed National Military Medical Center and University of Maryland - College Park

